# Genetic assessment of efficacy and safety profiles of coagulation cascade proteins identifies Factors II and XI as actionable anticoagulant targets

**DOI:** 10.1101/2023.12.05.23299567

**Authors:** Eloi Gagnon, Arnaud Girard, Jérôme Bourgault, Erik Abner, Estonian Biobank Research Team, Dipender Gill, Sébastien Thériault, Marie-Claude Vohl, André Tchernof, Tõnu Esko, Patrick Mathieu, Benoit J. Arsenault

## Abstract

**Background:** Anticoagulants are routinely used by millions of patients worldwide to prevent blood clots. Yet, problems with anticoagulant therapy remain, including a persistent and cumulative bleeding risk in patients undergoing prolonged anticoagulation. New safer anticoagulant targets are needed.

**Methods:** We performed two-sample Mendelian randomization (MR) and genetic colocalization to prioritize anticoagulant targets with the strongest efficacy (venous thromboembolism [VTE] prevention) and safety (low bleeding risk) profiles. We leveraged three large-scale plasma protein datasets (deCODE, n=35,559; Fenland n = 10,708; ARIC n= 7,213) and one liver gene expression dataset (n =246) to evaluate evidence for a causal effect of 26 coagulation cascade plasma proteins on VTE from a new genome-wide association meta-analysis of 44,232 VTE cases and 847,152 controls (from the UK Biobank, FinnGen and Estonian Biobank), stroke subtypes (from UK Biobank and International Stroke Genetics consortium 73,652 cases and 1,234,808 controls), bleeding outcomes (FinnGen, n=309,154) and over one million parental lifespans (UK Biobank and LifeGen consortium).

**Results:** Genetically predicted reductions in F2 blood levels were associated with lower VTE risk (OR [odds ratio] per 1 standard deviation [SD] lower F2=0.44, 95% CI=0.38-0.51, p=2.6E-28) and cardioembolic stroke risk (OR = 0.55, 95% CI=0.39-0.76, p=4.2e-04) but not with bleeding (OR = 1.13, 95% CI=0.93-1.36, p=2.2e-01). Genetically predicted F11 reduction were associated with lower risk of VTE (OR = 0.61, 95% CI=0.58-0.64, p=4.1e-85) and cardioembolic stroke (OR = 0.77, 95% CI=0.69-0.86, p=4.1e-06), but not with bleeding (OR = 1.01, 95% CI=0.95-1.08, p=7.5e-01) (Figure 3). These MR associations were concordant across the three blood protein datasets and the hepatic gene expression dataset as well as three different MR and colocalization analyses.

**Conclusion:** These results provide strong genetic evidence that F2 and F11 may represent safe and efficacious therapeutic targets to prevent VTE and cardioembolic strokes without substantially increasing bleeding risk.

## BACKGROUND

The coagulation system performs the delicate task of keeping blood in a fluid state while repairing damaged vessels. Too little coagulation and blood will leave the circulation system following an injury, causing bleeding. Too much and blood clots will block blood passage, causing thrombolytic events. Conditions involving thrombotic events (i.e. ischemic heart disease, stroke, and venous thromboembolism [VTE]) account for ∼2% of deaths worldwide, being among the leading cause of mortality (Lozano et al. 2012). As a result, anticoagulants are routinely used in millions of patients worldwide for the treatment of different thrombotic disorders (Wendelboe and Raskob 2016).

Yet, problems with anticoagulant therapy remain, including a persistent and cumulative bleeding risk in patients undergoing prolonged anticoagulation. Meta-analyses of clinical trials have reported up to a 11% higher frequency of major bleeding in patients on warfarin or non-vitamin K antagonist oral anticoagulants compared to placebo (Chai-Adisaksopha et al. 2014). Anticoagulant therapies with little to no bleeding risk remain out of reach, urging the medical community to identify new targets. All anticoagulant pharmaceutical therapies target one or more of the blood proteins involved in the coagulation cascade, many of which are synthesized by the liver. Optimal anticoagulant targets need to be safe (cause low bleeding risk) and efficacious (prevent thrombosis risk). New opportunities for unravelling coagulation targets with a favorable benefit/risk ratio arise from the widespread availability of gene expression, protein, and electronic health records data from hundreds of thousands of patients. These large-scale data can be connected using the Mendelian randomization (MR) analytic paradigm to proxy the lifelong consequences of genetic perturbations of drug targets (Daghlas and Gill 2023a).

Here, we perform two-sample (MR) to evaluate evidence for a causal effect of 26 coagulation cascade plasma proteins on VTE, stroke subtypes, bleeding outcomes and parental lifespan. We used blood protein quantitative trait loci (pQTLs) from three distinct genome-wide association studies (GWAS). We also leveraged liver expression quantitative trait loci (eQTLs) from a hepatic gene expression GWAS of 246 liver samples. These eQTLs and pQTLs were used as instruments to prioritize the therapeutic targets with the optimal efficacy (largest effect on VTE and stroke subtypes) and safety (lowest effect on bleeding outcomes) profiles.

## MATERIALS and METHODS

We performed colocalization analyses and two-sample MR investigation using GWAS summary-level data. There was minimal sample overlap between datasets. All participants were of European ancestry. We harmonized the exposure and outcome datasets by aligning the effect sizes of different studies on the same effect allele. When a SNP was not present in the outcome datasets, we used a proxy SNP (*r*^2^> 0.6) obtained using linkage disequilibrium matrix of European samples from the 1000 Genomes Project. All GWAS used are described in Supplementary Table 1.

### Study exposures

We included all proteins involved in the coagulation cascade as defined by PANTHER: a searchable database of gene products organized by biological function (Thomas et al. 2003), which were measured by the SomaScan version 4. A total of three blood pQTL datasets were used: deCODE, Atherosclerosis Risk in Communities (ARIC) and Fenland. In the deCODE population-based cohort, 4719 blood plasma protein levels were measured in 35,559 Icelanders with 4907 aptamers from SomaScan version 4 (Ferkingstad et al. 2021). The aptamer levels were adjusted for age and sex and the resulting residuals were inverse rank normal transform prior to GWAS. We extracted associations between 27 million SNPs measured by Illumina SNP chips and 26 blood coagulation proteins measured using 33 SOMAmers. In the ARIC population-based cohort, 4,657 plasma proteins were measured in 7,213 European Americans using 4,657 aptamers from SomaScan version 4 (Zhang et al. 2022). The relative abundance of SOMAmers was adjusted in a linear regression model including Probabilistic Estimation of Expression Residuals (PEER) factors and the covariates sex, age, study site, and 10 ancestry-based principal components. We extracted associations between cis-SNPs measured with Infinium Multi-Ethnic Global BeadChip array (Illumina, GenomeStudio) and 25 blood coagulation proteins measured using 32 SOMAmers. In this dataset, only summary statistics on cis-acting variants, defined as the transcription starting +/− 500Kb, are available. In the Fenland population-based cohort, 4,775 plasma proteins were measured in 10,708 European-descent participants with 4,978 apatamers using the SomaScan v4 assay (Pietzner et al. 2021). The aptamers levels were adjusted for age, sex, the first ten principal genetic components and test site and the resulting residuals were inverse rank normal transform prior to GWAS. We extracted association between SNPs and 26 blood coagulation proteins measured using 33 SOMAmers. We also used liver gene expression of coagulation cascade proteins as study exposures from the Institut Universitaire de Cardiologie et de Pneumologie de Québec (IUCPQ) Obesity Biobank (https://iucpq.qc.ca/en/research/platforms/biobank). Liver samples were obtained by incisional biopsy of left lobe and immediately snapped frozen. Read counts and TPM values were produced with RNA-SeQC v2.4.2 (DeLuca et al. 2012). Measures were adjusted for sex, age, the top 10 genetics principal components and 15 PEER factors. We mapped liver expression quantitative trait loci (eQTLs) using samples obtained from 246 participants that passed genotyping and RNA sequencing quality controls, as previously described (Gobeil et al. 2023).

### Study outcomes

VTE: We performed a GWAS meta-analysis of 3 cohorts: the UK Biobank, the Estonian Biobank, and FinnGen. We performed two novel GWAS for VTE in the Estonian Biobank and the UK Biobank (data application number 25205). In the UK Biobank and the Estonian Biobank, VTE cases were defined as self-reports confirmed by a trained medical professional of deep vein thrombosis, pulmonary embolism, or venous thromboembolism or according to EHR codes (ICD-10: I80.2, I80.2, I82.2, I26.0, and I26.9, OPCS-4: L79.1 and L90.2) (Klarin et al. 2017). Participants identified with cases of other forms of thrombosis (ICD-10: I81, I80.0, I80.3, I80.8, I80.9), known coagulation defects (D68) or Budd-Chiari syndrome (I82.0) were excluded from the analysis. Participants without these codes were identified as controls. We included a total of 17,200 European cases and 387,625 controls from the UK Biobank and 12,569 European cases and 164,827 control in the Estonian Biobank. We used the SAIGE (Scalable and Accurate Implementation of Generalized Mixed Models) (Zhou et al. 2018) algorithm to perform GWAS in the UK and Estonian Biobanks. This method adjusts for population-based factors such as sample relatedness while still maintaining maximum statistical power. We used sex, age, and the 10 first ancestry-based genetic principal components as covariates in the analysis. We then meta-analyzed these results with VTE GWAS summary statistics obtained from the FinnGen‘s data freeze 7 (Kurki et al. 2023). This GWAS included 14,454 cases and 387,625 controls. FinnGen GWAS includes summary statistics for 16 million genetic markers genotyped using the Illumina or Affymetrix arrays. The meta-analysis was completed using the METAL algorithm, using a fixed-effect inverse variance weighted meta-analysis (Willer, Li, and Abecasis 2010). Overall, the VTE GWAS meta-analysis includes 9,572,513 SNPs with minor allele frequency ≥0.01 in 44,223 cases and 847,152 controls. Stroke and stroke subtypes: We used GWAS summary statistics from a meta-analysis of the International Stroke Genetics Consortium (ISGC) consortium and the UK Biobank totalling 73,652 cases of stroke 1,234,808 controls of European ancestry (Mishra et al. 2022). Cases were defined with clinical diagnosis of strokes (62,100 cases of ischemic stroke; 10,804 cases of cardioembolic stroke; 6,399 cases of large artery stroke; 6,811 cases of small vessel stroke; and 1,234,808 controls). The GWAS were adjusted for age, sex, principal components of population stratification and study-specific covariates when needed. Bleeding: For bleeding outcomes, we used GWAS summary statistics from a GWAS of the FinnGen population based cohort totalling >309,000 individuals of European ancestry (Kurki et al. 2023). Cases were established with electronic health record ICD10 codes (GI bleeding ICD10 = K92[0-2]; injury to the elbow and forearm ICD10 = S50, intracranial haemorrhage ICD10 = “I9_SAH|I9_ICH”; bleeding ICD10 = “D3_HAEMORRHAGCIRGUANTICO|H7_CONJUHAEMOR|H7_RETINAHAEMOR R|H7_VITRHAEMORR|H7_CHORHAEMORRHAGE|ST19_EPIDU_HAEMORRHAG E|I9_INTRACRA|I9_OTHINTRACRA|ST19_TRAUMAT_SUBDU_HAEMORRHAGE |ST19_TRAUMAT_SUBAR_HAEMORRHAGE|R18_HAEMORRHAGE_RESPI_PAS SA|K11_GIBLEEDING”,). The GWAS was performed in SAIGE (v.0.35.8.8) and adjusted for sex, age, genotyping batch and ten first principal genetic components as covariates. Parental Lifespan: We used GWAS summary statistics for two parents’ survival from a meta-analysis of the UK Biobank and the LifeGen consortium of 26 population cohorts (n=1,012,240; all of European ancestry) (Timmers et al. 2019). The outcome was defined as parental survival in a Cox model. Mother and father survival information was combined assuming the effects were the same in men and women.

### Mendelian randomization

We applied three types of MR analysis to assess the causal effect of coagulation factors on VTE, stroke, bleeding outcomes and parental lifespan. Cis-acting pQTLs (close to the gene of interest) are more specific instruments, suppressing or upregulating the expression of a gene, whereas trans pQTLs (distal to the gene) may operate via more complex mechanisms, making them more likely to be pleiotropic. First, we used a uni-cis-MR framework. We selected the top SNP with the smallest p-value in a 1 Mb window of the coagulation cascade gene regions. MR estimates on each protein and outcomes were obtained with the Wald ratio. The Wald ratio is calculated by dividing the SNP-outcome effect by the SNP-exposure effect. To assess instrument strength, we used the F-statistic (Burgess, Thompson, and CRP CHD Genetics Collaboration 2011), and to quantify the variance explained, we used the *r^2^* value (Pierce, Ahsan, and VanderWeele 2011). An issue that can arise when performing uni-cis MR analysis is the potential inclusion of functional variants altering the structure of the protein and creating epitope binding artifacts. To evaluate the possibility of epitope binding artifacts, we annotated all genetic instruments using the variant effect predictor and removed instruments with evidence of being altering variant. We evaluated reverse causality by performing the Steiger test. The Steiger test provides a p-value under the null hypothesis that the difference in variance explained is null (Hemani, Tilling, and Davey Smith 2017). We performed the Steiger test to identify variants with evidence of a stronger association with the outcome than with the exposure.

We also used a multi-cis MR approach where we selected multiple independent cis-acting variants as genetic instruments. Multi-cis MR analysis, that is, using multiple genetic variants allows performing robust MR analyses to evaluate robustness to horizontal pleiotropy in the causal estimates (Gkatzionis et al., 2021). For this analysis, we included as genetic instruments all cis-acting SNP in a 1MB window around the gene region independently (LD clumping = r^2^ <0.1) associated at p <5e-08 with protein levels. Third, we used pan MR analysis (all cis- and trans-acting variants). We selected independent (r^2^<0.01) genome-wide significant SNPs (p <5e-08) from all regions of the genome and performed MR analysis. The inclusion of trans-acting pQTLs increases the number of independent genetic instruments, thus increasing power, allowing the use of robust analysis and the quantification of heterogeneity (Cochran’s Q) to assess the validity of the MR assumptions, but can introduce pleiotropy.

For multicis and pan-MR analysis, we performed the inverse variance weighted (IVW) method with multiplicative random effects with a standard error correction for under dispersion as primary MR analysis (Burgess, Foley, and Zuber 2018). We also perform three different robust MR analyses (the MR Egger (Bowden, Davey Smith, and Burgess 2015), the contamination mixture (Burgess et al. 2020), the weighted median, and the MR PRESSO (Verbanck et al. 2018) to explore if their estimate substantially differed from the primary MR results. These methods, their assumptions and their statistical property have been extensively reviewed elsewhere (Slob and Burgess 2020).

### Genetic colocalization

Spurious association can occur because of linkage disequilibrium (LD). LD bias occurs when the true causal SNP of the exposure and the outcomes are distinct, but in LD, causing a false positive MR association. To evaluate bias due to linkage disequilibrium, we performed genetic colocalization analysis. We evaluated the posterior probability that both the protein or the RNA and the outcomes shared a single variant using a Bayesian model implemented in the *coloc* R package (Wallace 2020). We included all variants in a 1MB window of the gene regions. We used the default priors for the analysis. We used a posterior probability H4 > 0.50 as a threshold for evidence of colocalization signifying that colocalisation is more likely than any other scenario combined (Cronjé et al. 2023).

### Code and data availability

Code for this manuscript can be found at https://github.com/gagelo01/F2_F11. The GWAS summary statistics for VTE will be deposited on the GWAS catalog upon acceptation of the manuscript. All other genome-wide summary statistics used in this study are in the public domain.

## RESULTS

### Venous thromboembolism genome-wide association meta-analysis

We performed a new VTE GWAS meta-analysis including two new GWAS in the Estonian Biobank and UK Biobank and an existing GWAS from FinnGen. The meta-analysis includes 44,223 cases identified through EHRs and self-identifications confirmed by trained nurses and 847,152 controls. This meta-analysis identified 3,017 single nucleotide polymorphisms (SNPs) significantly associated with VTE (p<5e-08) spread throughout the genome at 77 independent risk loci (LD *r^2^*<0.01)(Supplementary Figure 1 and Supplementary Figure 2), all of which have been previously identified (Ghouse et al. 2023)

### Estimated effects of plasma coagulation proteins using uni-cis, multi-cis and pan MR

As discovery methods, we first performed uni-cis MR where we used the variant with the lowest p-value in the cis-acting region ±1MB as instruments and calculated the effect with a Wald ratio. Most proteins were measured in the three cohorts (deCODE, ARIC, and Fenland). We used the deCODE dataset for discovery analysis and the two other datasets as replication analyses. Across all study cohorts, all proteins had at least one top SNP with a F statistic >10 indicating sufficient strength of instruments. The top SNPs of the PLAT, PLAUR, and KNG1genes were variant altering the amino acids sequence. Epitope binding artifact was unlikely for the remaining 23 proteins. To evaluate reverse causality, we performed the Steiger test, where genetic instruments that explain more variance in the outcome than the exposure are tagged. No genetic instruments were tagged by Steiger test indicating that reverse causality was unlikely.

Using the uni-cis MR framework, among all the 26 coagulation cascade blood proteins studied, F2 and F11 most strongly affected VTE and ischemic stroke (Figure 1). In deCODE, genetically predicted reductions in F2 blood levels (mimicking the effect of F2 inhibitors) as measured with the aptamer 5316_54 was associated with lower VTE risk (OR [odds ratio] per 1 standard deviation [SD] lower F2=0.44, 95% CI=0.38-0.51, p=2.6E-28) and cardioembolic stroke (OR = 0.55, 95% CI=0.39-0.76, p=4.2e-04), with a comparatively smaller increased risk in bleeding (OR=1.13 95%, CI=0.93-1.36, p=2.2e-01). Similarly, genetically predicted reductions in F11 blood levels (mimicking the effect of F11 inhibitors) were associated with lower risk of VTE (OR=0.61, 95% CI=0.58-0.64, p=4.1e-85) and cardioembolic stroke (OR = 0.77, 95% CI=0.69-0.86, p=4.1e-06), but were not associated with bleeding (OR=1.01, 95% CI=0.95-1.08, p=7.5e-01) (Figure 2). MR results were similar in ARIC and Fenland datasets (Supplementary Figure 3).

**Figure 1.**
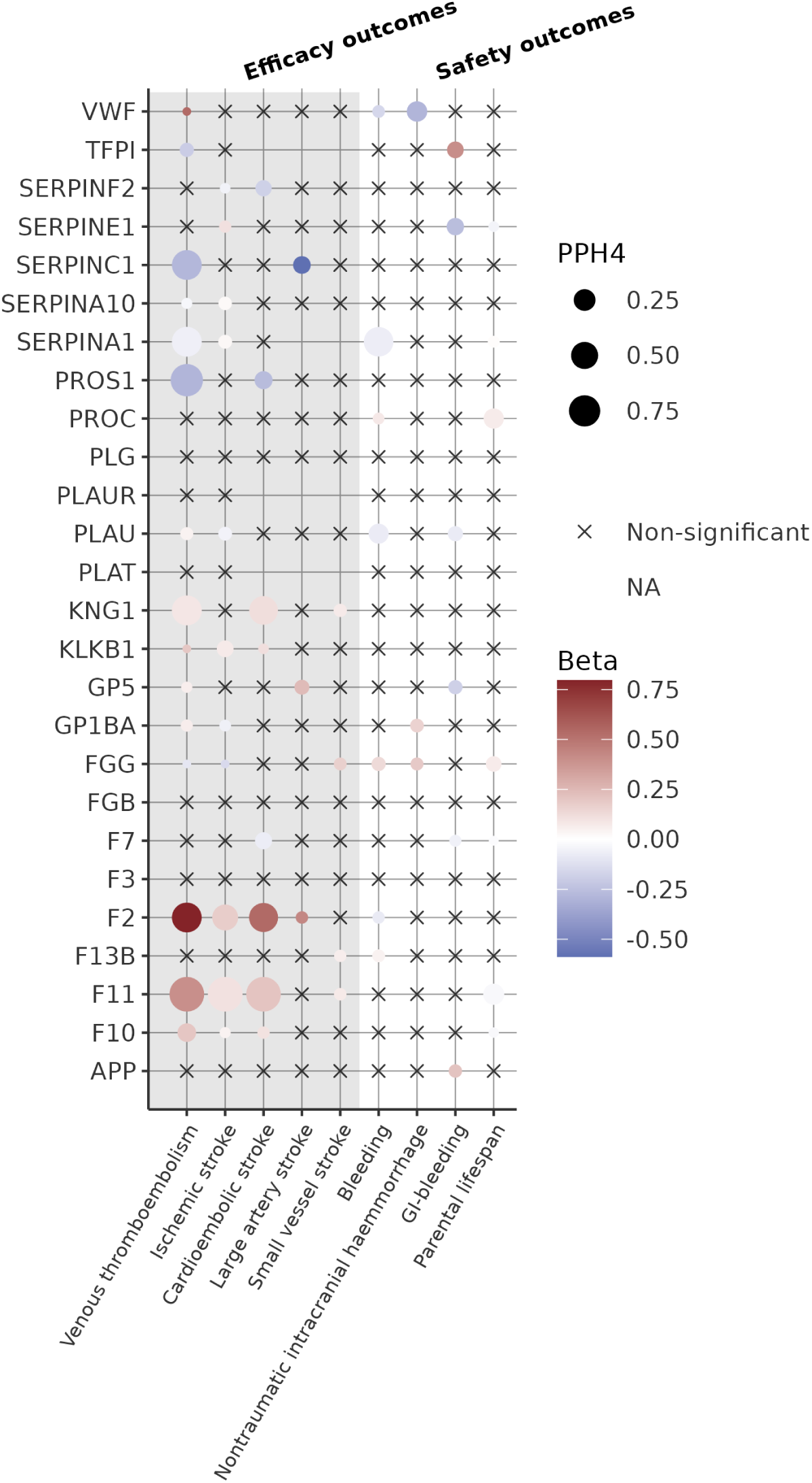
Genetically predicted effect of 26 coagulation pathway blood protein level on safety and efficacy outcomes. Betas and PPH4 are averaged across the three study cohorts deCODE, Fenland and ARIC. Non-available (NA) associations stem from a lack of overlapping SNPs or proxies (r^2^>0.6) between exposure and outcome data resulting in no overlapping SNP in the harmonized data set. Associations at p-value >0.05 are depicted with crosses. The mean PPH4 of colocalization is depicted with the size of the balloon.

**Figure 2.**
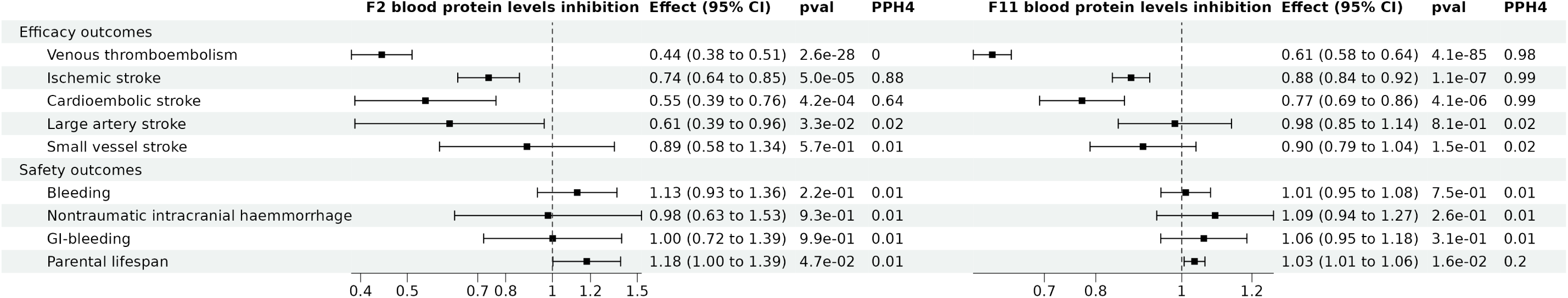
Genetically predicted reductions in blood F2 and F11 levels on safety and efficacy outcomes in a uni-cis MR analysis. Effect of 1 standard deviation decrease (inhibition) in F2 or F11 blood protein levels in deCODE on efficacy and safety outcomes. PPH4 stands for “posterior probability of hypothesis 4” as calculated with the coloc method, the posterior probability that two traits share a common variant.

Bayesian genetic colocalization revealed strong probability of shared variants between F2/F11 and VTE/cardioembolic stroke, indicating that confounding by LD was unlikely. Across samples and aptamers, the mean PPH4 for F2 and VTE was 0.66, for F2 and cardioembolic stroke was 0.61, for F11 and VTE was 0.99; and for F11 and cardioembolic stroke was 0.99. There was no evidence of colocalisation between F2 and bleeding and F11 and bleeding (PPH4<0.02) (Supplementary Figure 4, supplementary table 2).

Multi-cis MR analyses provided concordant evidence that F2 and F11 inhibition was associated with decreased risk of VTE and stroke and not associated with bleeding (Figure 3). For multi-cis MR, we selected all genome-wide significant SNPs in the cis region ± 1MB and pruned them with a correlation threshold of *r^2^*<0.1. Consistent with what was obtained using the uni-cis MR analysis, genetically predicted reductions in F2 blood levels were associated with decreased VTE risk (OR = 0.46, 95% CI=0.41-0.52, p=3.5e-33) and cardioembolic stroke risk (OR = 0.58, 95% CI=0.44-0.78, p=2.8e-04), with a comparatively smaller increase risk in bleeding (OR = 1.14, 95% CI=0.99-1.31, p=6.2e-02). Similarly, genetically predicted F11 reduction were associated with lower risk of VTE (OR = 0.66, 95% CI=0.63-0.69, p=3.9e-64) and cardioembolic stroke (OR = 0.84, 95% CI=0.79-0.88, p=8.5e-10), but not with bleeding (OR = 0.99, 95% CI=0.96-1.02, p=6.5e-01) (Figure 3). MR results were similar using robust MR analyses and similar in ARIC and Fenland datasets (Supplementary Figure 5, Supplementary Table 3). Altogether, multi-cis MR provides convergent evidence that F2 and F11 inhibition is associated with a decrease in VTE and cardioembolic stroke risk, but not associated with bleeding risk.

**Figure 3.**
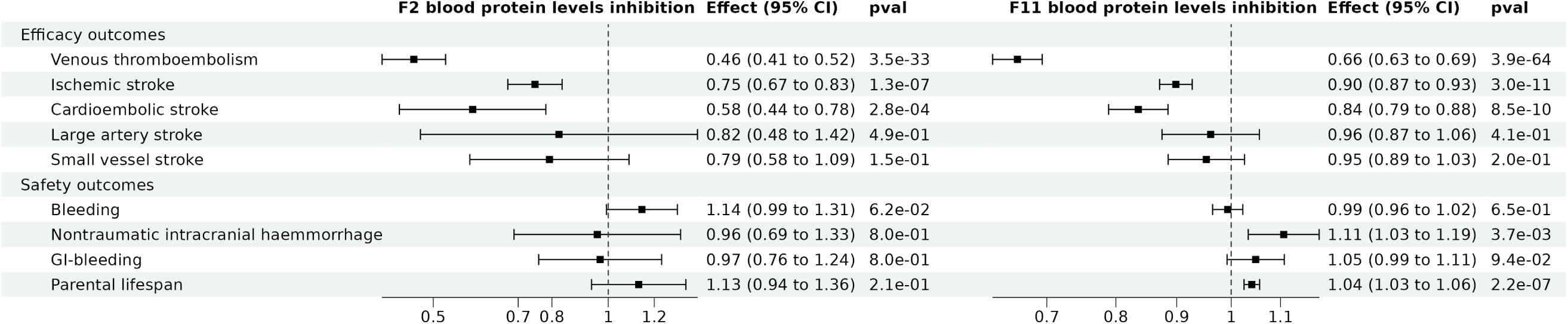
Genetically predicted reductions in blood F2 and F11 levels on safety and efficacy outcomes in a multicis MR analysis. Effect of 1 standard deviation decrease (inhibition) in F2 or F11 blood protein levels measured in deCODE on efficacy and safety outcomes. PPH4 stands for “posterior probability of hypothesis 4” as calculated with the coloc method, the posterior probability that two traits share a common variant.

Pan MR analysis revealed a strong effect of F2 and F11 on VTE and cardioembolic stroke (Figure 4). Trans-acting pQTLs are more likely to act on protein levels via pleiotropic pathways. However, their inclusion in MR analysis can increase the variance explained and prevent reliance on a single genetic region. We therefore included all genome-wide significant SNPs throughout the genome clumped at r^2^<0.01 in a 1 MB window region and performed a pan MR analysis. The inverse variance weighted method was used as primary MR analysis and other robust MR analyses (MR PRESSO, contamination mixture, weighted median) were used as sensitivity analyses. In the deCODE sample, using pan MR analyses, genetically predicted reduction in F2 blood levels was associated with lower VTE risk (OR = 0.56, 95% CI=0.42-0.75, p=9.5e-05) and cardioembolic stroke (OR = 0.52, 95% CI=0.40-0.67, p=2.7e-07) risk, but did not associate with bleeding (OR = 0.98, 95% CI=0.89-1.08, p=6.9e-01). Similarly, genetically predicted F11 reduction was associated with lower risk of VTE (OR = 0.79, 95% CI=0.74-0.84, p=7.9e-13) and cardioembolic stroke (OR = 0.86, 95% CI=0.81-0.91, p=4.9e-07), but not bleeding (OR = 1.00, 95% CI=0.96-1.03, p=9.0e-01) (Supplementary Table 4). For F11, these effects were significant and directionally consistent across all robust MR methods indicating robustness to pleiotropy and replicated in the Fenland cohort (Figure 4). For the association between F2 and VTE, the contamination mixture method did not reach significance in deCODE, but did in Fenland. For the effect of F2 on VTE and cardioembolic stroke, several robust MR methods did not reach statistical significance for the aptamer 4157_2, but did with the aptamer 5316_54. Violation of the exclusion restriction assumption therefore cannot be ruled out for the aptamer 4157_2. We could not perform pan-MR analysis in the ARIC cohort, because only cis summary statistics are publicly available.

**Figure 4.**
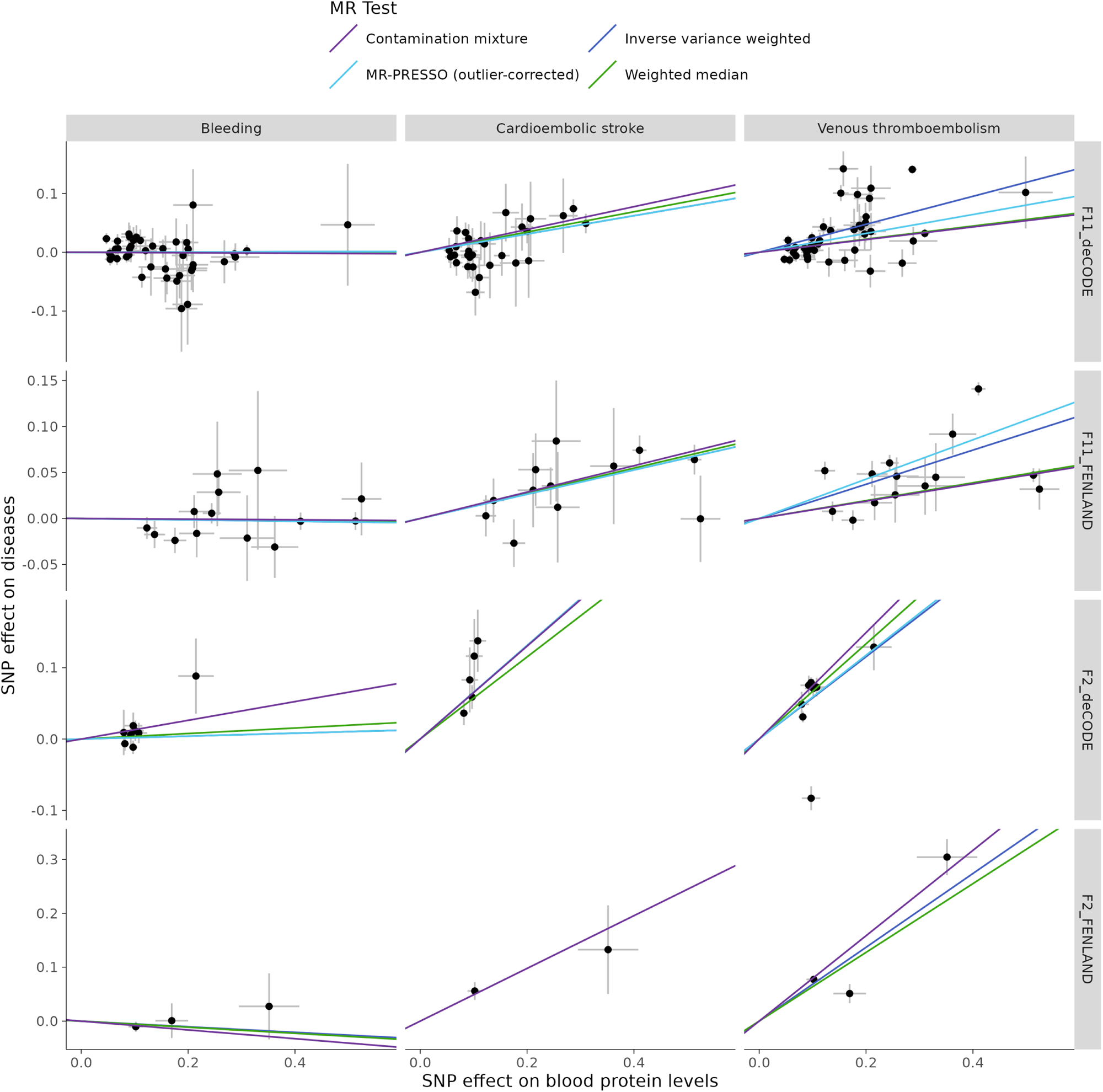
Genetically predicted reductions in blood F2 and F11 levels on safety and efficacy outcomes in a pan MR analysis. Scatter plot of the association between F2 and F11 in deCODE sample on ischemic stroke, venous thromboembolism, and bleeding. Inverse variance weighted methods and robust MR analyses are presented.

**Figure 5.**
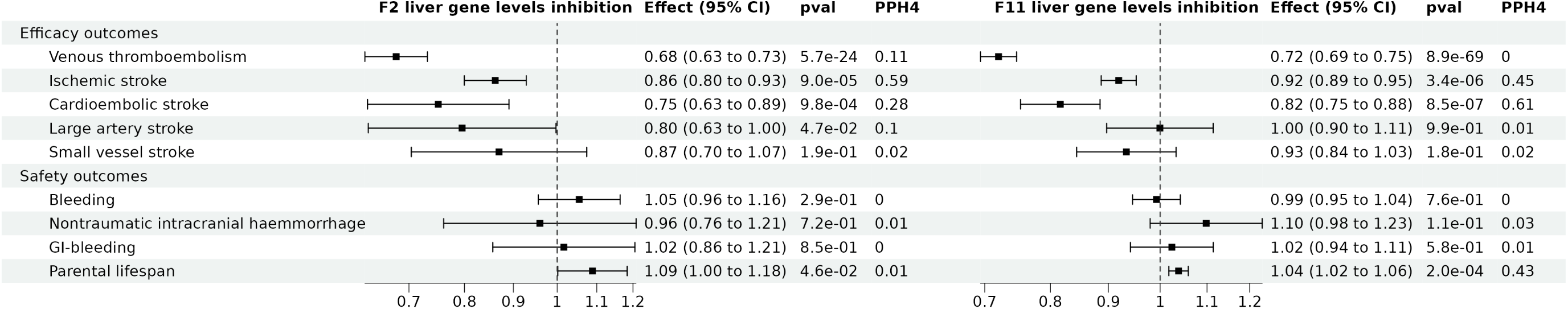
Genetically predicted reductions in liver expression of *F2* and *F11* on safety and efficacy outcomes in a uni-cis MR analysis. Effect of 1 standard deviation decrease (inhibition) in F2 or F11 hepatic gene expression levels on efficacy and safety outcomes. PPH4 stands for “posterior probability of hypothesis 4” as calculated with the coloc method, the posterior probability that two traits share a common variant.

### Impact of liver gene levels of F2 and F11 on efficacy and safety outcomes

F2 and F11 are specifically expressed in the liver and are then secreted in the bloodstream. We tested the hypothesis that hepatic expression of F2 and F11 might also be causally associated with efficacy and safety outcomes. To test this hypothesis, we used the same uni-cis MR approach as with the blood proteins, but used instead hepatic gene expression levels (methods). Because the sample size was lower for this dataset, the top SNPs had larger p-value (1.1e-05 for F2 and 3.0e-12 for F11). Variance explained was nevertheless high (*r^2^* = 0.08 for *F2* and 0.18 for *F11*) and the instrument was strong (F-statistic of 20 for *F2* and 55 for *F11*). Using the Wald ratio formula, genetically predicted reduction in *F2* hepatic gene level was associated with lower VTE risk (OR per 1 SD lower *F2*=0.68, 95% CI=0.63-0.73, p=5.7e-24), lower cardioembolic stroke risk (OR = 0.75, 95% CI=0.63-0.89, p=9.8e-04), and increased lifespan (0.08 years 95% CI=0.00-0.17, p=4.6e-02), but did not associate with bleeding (OR=1.05, 95% CI=0.96-1.16, p=2.9e-01). Genetically predicted reductions in liver *F11* gene levels were associated with lower VTE risk (OR=0.72, 95% CI=0.69-0.75, p=8.9e-69), lower cardioembolic stroke risk (OR = 0.82, 95% CI=0.75-0.88, p=8.5e-07), and increased lifespan (0.04 years 95% CI=0.02-0.06, p=2.0e-04), but did not associate with bleeding (OR=0.99, 95% CI=0.95-1.04, p=7.6e-01) nor any other bleeding outcomes. Overall, the effect size of hepatic gene expression was smaller than that of blood proteins. This is expected since blood protein levels is a more proximal effector of blood coagulation and presumably not all hepatic mRNA is transcribed into proteins.

Colocalisation yielded low-to-moderate probability of shared causal variant between F2/F11 and VTE/ischemic stroke. The PPH4 for F2 and VTE was 0.11, for F2 and cardioembolic stroke was 0.28, for F11 and VTE was 0.00; and for F11 and cardioembolic stroke was 0.61 (Supplementary Figure 4). The colocalization between liver F11 and VTE yielded a PPH4 of 0 and a PPH3 of 0.99, contrasting with the colocalization between blood F11 protein levels and VTE which yielded a PPH4 of 0.99 and a PPH3 of 0 (Supplementary Table 2). The difference in colocalisation results is most likely due to differential sample size. Overall, these analyses support that reduction of hepatic gene levels of F2 and F11 are associated with lower VTE and ischemic stroke, but not bleeding risk.

## DISCUSSION

To prioritize new targets for anticoagulant therapy, we performed a new VTE GWAS and examined evidence for a causal effect of 26 coagulation cascade plasma proteins on VTE, stroke subtypes, bleeding outcomes and parental lifespan. Using a comprehensive MR strategy, we found that genetically predicted inhibition of F2 and F11 were associated with lower risk of VTE and ischemic stroke, without substantially increasing bleeding risk. These associations were concordant across three large blood pQTL datasets, one liver eQTL dataset and across several robust MR methods. Altogether, if substantiated by randomized clinical trials, these results provide strong genetic evidence that F2 and F11 may represent safe and efficacious therapeutic targets to prevent VTE and cardioembolic stroke without substantial increase in bleeding risk.

Our MR results on F2 are concordant with results of clinical trials comparing direct thrombin inhibitors with placebo (Schulman et al. 2013; Devereaux et al. 2018). Direct thrombin inhibitors specifically inhibit the intrinsic activity of the F2 gene product: the protein thrombin. The administration of the direct thrombin inhibitors dabigatran at a dose of 150 mg twice daily strongly reduced the risk of venous thromboembolism (hazard ratio, 0.08; 95% CI 0.02 to 0.25), but increased the risk of major or clinically relevant bleeding (hazard ratio, 2.92; 95% CI 1.52 to 5.60) when compared to placebo in patients with venous thromboembolism (Schulman et al. 2013). Similarly, Dabigatran 110 mg versus placebo twice daily lowered the 2-year risk of major vascular complications (hazard ratio, 0·72 95% CI 0.55 to 0.93), but increased risk of minor bleeding (hazard ratio, 1.64 95% CI 1.25 to 2.15), but not major bleeding, among patients who had myocardial injury after non-cardiac surgery (Devereaux et al. 2018). Our MR results similarly outline this advantageous efficacy/safety ratio.

Preclinical and clinical investigations support a role of *F11* inhibition in decreasing VTE risk with mild increase in bleeding risk. First, *F11*-deficient mice do not appear to have hemostatic defects. Indeed, bleeding time in mice lacking *F11 is* comparable to wild type mice after injury to the tail (Ay et al. 2017; Gailani, Lasky, and Broze 1997; Wang et al. 2005). One study, however, found that *F11*-deficient mice display a moderate hemostatic defect in a saphenous vein bleeding model (Ay et al. 2017). But this result failed to replicate in another study where saphenous vein bleeding time was the same in *F11^−/−^* mice compared to wild-type mice (Mohammed et al. 2018). Second, patients with severe *F11* deficiency have a 4.68 lower risk of VTE, a 8.56 lower risk of ischemic stroke, but the same incidence of myocardial infarction compared to individuals with normal *F11* function (Salomon et al. 2011; 2008). *F11*-deficient individuals are not at higher risk of spontaneous bleeding, but risk of bleeding following trauma or surgery increases (Bolton-Maggs 2009). Third, a previous MR investigation, using a different instrument selection strategy, provided evidence that lower levels of F11 protect against VTE and cardioembolic stroke, but does not increase bleeding risk (Daghlas and Gill 2023b). The latter study, however, did not test other coagulation cascade proteins or liver gene expression, so they could not prioritize targets or assess the effect of perturbing liver gene expression. Finally, several types of *F11* inhibitors are currently being tested in humans and have demonstrated good efficacy and safety. Published phase 1 trials do not report any safety concerns for *F11* inhibitors (Poenou et al. 2022). F11 inhibition with liver-targeted antisense oligonucleotides (*F11*-ASO) (Büller et al. 2015), small molecules (milvexian)(Weitz et al. 2021) or monoclonal antibodies (abelacimab) (Verhamme et al. 2021) s in patients undergoing total knee surgery were significantly superior to enoxaparin, in terms of rates of VTE.

RNA therapeutics targeting hepatic *F2* or *F11* are promising anticoagulant targets. RNA therapeutics such as ASOs or small interfering RNAs are used to silence gene expression by complementary base pairing with messenger RNA. The liver is an organ of choice for ASO delivery. ASOs have been chemically engineered to bind asialoglycoprotein receptors, which are present on the surface of hepatocytes, but not on other cell types or tissues. ASOs for *F11* mRNA are currently under development (Büller et al. 2015). In the *F11*-ASO TKA study, 300 patients undergoing total knee surgery were randomized to receive one of two doses of an ASO against *F11* (IONIS F11-LRx 200 mg or 300 mg) or 40 mg of enoxaparin once daily (Büller et al. 2015). At 200-mg, *F11*-ASO was non-inferior (27%), while the 300-mg was superior (4%) to enoxaparin (30%) (P<0.001) for preventing a composite outcome of symptomatic VTE or asymptomatic deep venous thromboembolism. Rates of bleeding were inferior in the *F11*-ASO groups (both 3%) than in the enoxaparin group (8%) (Büller et al. 2015). ASO targeting possesses a convenient mode of delivery, an efficient reversal strategy, and good efficacy (prevent thrombosis) and safety (low bleeding risk) profile. ASO therapy decreases *F11* levels so it could be rapidly reversed with an injection of *F11*. Although ASO therapies are currently only delivered using subcutaneous injection, there long half-life allows conveniently for weekly or potentially monthly self-administration. Our results from genetically predicted liver expression of the genes encoding *F2* and *F11* support further investigation into the role of RNA interfering therapies in the prevention of VTE.

The main strength of this study is the triangulation of evidence using several datasets and several MR methodologies providing convergent results. The main limitation of the study is that all anticoagulant proteins could not be evaluated. There are at least 40 proteins involved in the coagulation pathway (Thomas et al. 2003), but the Somascan V4 only assesses 26 of them. It is possible that other coagulation cascade proteins may have more optimal efficacy or safety profile, but they could not be tested in our MR framework.

In conclusion, these results support that inhibition of hepatic *F2* and *F11* genetic expression and/or blood protein levels may reduce the risk of thrombosis without substantially increasing the risk of bleeding. *F2* is an established target for anticoagulation. *F11* is an emerging target of three different anticoagulant therapies under development with various inhibition strategies. These findings provide genetic support for therapies either targeting hepatic gene expression or blood protein levels of *F2* or *F11*, potentially opening a new era of anticoagulant therapies.

## Data Availability

The GWAS summary statistics for VTE will be deposited on the GWAS catalog upon acceptation of the manuscript. All other genome-wide summary statistics used in this study are in the public domain.

## ACKNOWLEDGEMENTS

We would like to thank all study participants as well as all investigators of the studies that were used throughout the course of this investigation. EG holds a doctoral research award from the *Fonds de recherche du Québec: Santé* (FRQS). JB both hold a master research award from the FRQS. BJA holds a senior scholar award from the FRQS. MCV is Canada Research Chair in Genomics Applied to Nutrition and Metabolic Health.

